# Determining the variability associated with measuring visual acuity and refractive error measurements: a systematic review

**DOI:** 10.1101/2025.01.17.25320329

**Authors:** C Van der Zee, MB Muijzer, JLJ Claessens, RPL Wisse

## Abstract

**Topic:** This systematic review examines variability regarding measuring visual acuity (VA) and refractive error (RE).

**Clinical Relevance:** VA and RE are key factors that determine visual function, and although accurately measuring VA and RE is fundamental to clinical practice, no scientific consensus has been reached with respect to variability. This lack of consensus affects our ability to establish a limit of agreement (LoA) for clinically accepted variability and may subsequently affect the clinical decision-making process, regulations, and research in the field of ophthalmology.

**Methods:** This systematic review was performed in accordance with PRISMA guidelines and was registered at PROSPERO (ID: CRD42024554663). We searched the Medline, PubMed, and Embase databases. The Quality Assessment of Diagnostic Accuracy Studies 2 (QUADAS-2) tool was then used to assess the risk of bias (RoB). We included studies that directly compared the accuracy of VA or RE and reported the LoA and mean difference expressed in logarithm of the minimum angle of resolution (LogMAR) for VA and diopters (D) for refraction. We then report the summarized mean LoA values with 95% confidence intervals (CIs).

**Results:** After applying the inclusion and exclusion criteria, our analysis included 13 and 6 studies that reported VA and RE measurements, respectively. From these 19 studies, only 6 (32%) scored a low RoB in at least three out of four QUADAS-2 domains. In 27 out of 38 subgroups (71%), LoA exceeded the generally accepted ranges for VA or RE (namely, ±0.15 LogMAR and ±0.5D, respectively). Based on our analysis, the summarized mean LoA values were ±0.20 LogMAR (95% CI: 0.17, 0.23) and ±0.70D (95% CI: 0.50, 0.89) for VA and RE measurements, respectively.

**Conclusions:** We found that both VA and RE measurements have relatively high variability, exceeding currently accepted limits. Thus, the currently accepted limits of this variability are either unrealistic or reflect studies with methodological weaknesses. Rigorous studies are therefore needed in order to more accurately estimate the acceptable limits of variability for both VA and RE measurements. For now, we propose using the mean LoA values obtained in our study as a provisional frame of reference. In addition, we discuss several aspects of understanding the variability that exists when measuring VA and RE, particularly with respect to comparing studies.

## Introduction

In the mid-19^th^ century, Dutch physiologist Prof. F.C. Donders published his dissertation regarding the refractive states of the eye, while Prof. H. Snellen Sr. reported a method to quantify visual acuity (VA) using optotypes; today, measuring VA and refractive error (RE) are routinely performed by eye care professionals.^1,2^ Indeed, both VA and RE are now considered trusted parameters that often guide clinical treatment decisions designed to improve the patient’s visual quality of life.^3^

The ability to reliably and accurately measure VA and RE is highly relevant to clinicians and patients alike, but also for clinical studies, screening initiatives, therapies, registries,^4^ healthcare insurers,^5^ and summative visual assessments (for example, to obtain a driver’s license^6^ or qualify for military service^7^). These parameters are also relevant globally, as at least 2.2 billion people worldwide currently have impaired near vision or distance vision, with refractive error being the leading cause. Moreover, studies estimate that by the year 2050 a striking half of the world’s population will be near-sighted.^8^ Together, these visual impairments are associate with an enormous financial burden, with global annual productivity costs estimated at approximately $411 billion.^3^ Thus, understanding better the accuracy and variability of measuring both VA and RE is crucial for improving the clinical decision-making process, developing suitable guidelines and regulations, screening patients, and designing research in the field of ophthalmology.

Despite its high clinical relevance, variability with respect to measuring VA and RE has not been reviewed previously. In addition, this variability can be affected by a number of biases. For example, these measurements are psychometric in nature, as the results depend in part on how the patient responds during the test. Moreover, a certain degree of variability is inevitable when comparing different optotypes (e.g., Sloan letters vs. Tumbling E) due to the so-called conversion effect.^9,10^ In addition, variability when measuring VA and RE is higher in patients with limited VA, thus compounding the importance of accurately measuring VA in order to guide the treatment process. Despite its widespread use and importance in decision making, a growing body of evidence indicates that the reported variations in measuring VA and RE may be higher than commonly accepted limits, making these conventions less reliable than consensus suggests. To address this possibility, we systematically reviewed studies that reported variability in measuring VA and RE, with the aim of establishing a new consensus in order to improve scientific, clinical, and regulatory decision-making processes. An additional aim of this review is to better explain this variability to the general reader in order to help guide the clinical decision-making process. Finally, we provide a report regarding the nature of the variability in measuring VA and RE in the adult population, and we reflect on how this variability aligns with current practice standards.

## Methods

### Protocol and registration

The protocol for this systematic review was designed in accordance with the guidelines outlined in the Preferred Reporting Items of Systematic Reviews and Meta-Analyses (PRISMA) statement.^11^ This systematic review was registered in PROSPERO on 15/06/2024 (ID: CRD42024554663).

### Eligibility criteria

We use strict inclusion criteria, although no date range was applied to the search. We included studies that involved adult participants (≥18 years of age) and used the following methods (or combination thereof):

- For measuring VA: Early Treatment Diabetic Retinopathy Study (ETDRS), Lovie-Bailey, Snellen, Landolt C, and/or Tumbling E charts.
- For measuring RE: Two measurements of manifest refraction (MR) were compared, excluding studies that used autorefraction.

In our search, for studies that investigated VA testing methods no restrictions were placed on ocular health, whereas for RE we included only studies that involved healthy subjects. As noted above, for RE we excluded studies that used autorefraction. We also excluded studies that lacked the following: the relevant outcome measurement (i.e., LoA or the mean difference with standard deviation), availability of the full-text manuscript, or clinical measurements (e.g., studies that predicted outcome and meta-analyses); we also excluded studies that used telemedicine, as well as conference proceedings, editorials, letter to the editor, and studies involving refractive surgery research or intraocular lens research. With respect to VA specifically, we excluded studies that did not report VA in Snellen or LogMAR, studies that did not measure distance visual acuity (DVA), and studies in which DVA was <1.3 LogMAR (corresponding to 0.05 decimal or 20/400 Snellen).

The gold standard for assessing agreement between two measurements is to perform a Bland-Altman analysis.^12^ In this analysis, two concepts are reported, namely structural bias (i.e., the mean difference) and random error (i.e., the range of differences expressed as the 95% limits of agreement, or LoA). Because these concepts are relevant to our analysis, they are defined in Textbox 1. In brief, structural bias refers to the systematic difference between two measurements, whereas random error refers to the interval within which 95% of all measurements are expected to fall. The level of agreement between two methods at a population level (i.e., the variability) can be combined with a context-specific understanding of the degree of variability considered to be relevant and acceptable. Relevant outcomes include a measure of repeatability and reproducibility in the VA (in LogMAR) and RE (in diopters, D) readings, defined as the structural bias in mean difference and random error in 95% LoA. Historically, the commonly acceptable limits defined in the literature are ±0.15 LogMAR and ±0.5D for VA and RE, respectively.^13–15^

### Search strategy for identifying studies

We performed a systematic literature search in accordance with PRISMA guidelines and the standards for formatting a systematic review. This search was performed on May 15, 2024, and included the PubMed, Medline, and Embase databases. Consideration was given to selecting search terms, languages, titles, abstract, keywords, MeSH Terms, and Emtree terms. Included terms were based on the population (adults, adulthood, aged, middle-aged, elderly), the visual acuity intervention (visual acuity charts, visual acuity test, ETDRS, Snellen, Landolt-C, Tumbling-E), and the refraction intervention (Refraction, manifest refraction, spherical equivalent). A list of the search terms, derivatives and synonyms is provided in Supplementary Table S1. During the full-text assessment, additional relevant studies that were not retrieved by the initial search were identified using snowballing and cross-referencing.

### Study selection

Two reviewers (authors C.Z. and M.M.) independently screened the records in a blinded fashion using the Rayyan platform. Any differences of opinion were resolved by consensus or by discussion with a third reviewer (author R.W.). Duplicate studies were manually reviewed before exclusion.

### Data collection and risk of bias assessment

Study data were extracted independently by two reviewers (authors C.Z. and M.M.), and each defined study population was included as a separate input. The following data were extracted from each study: the number of subjects (n), age (in years), method(s) compared, mean difference, SD, and/or 95% LoA. The risk of bias (RoB) in each study was assessed independently by two reviewers (authors C.Z. and M.M.) using the Quality Assessment of Diagnostic Accuracy Studies 2 (QUADAS-2) tool,^16^ and any disagreements were resolved through consensus. QUADAS-2 consists of four key domains: patient selection, index test, reference standard, and flow & timing. The tool and signalling questions were tailored to address our review’s research question. In addition to the regular signaling questions, we assigned a higher RoB to studies in which the observer was not blinded, the reference and index tests were not performed in a random sequence, the testing protocol was not described, an ETDRS chart was not used as a reference standard (in case of non–test-retest studies), and/or the interval between the reference and index tests was longer than 1 day.

### Data synthesis and analysis

All extracted results were transformed to the appropriate format (e.g., to LogMAR notation in the case of VA). If not reported in a study, if possible missing results were calculated from the reported data (e.g., by calculating LoA from the SD). Based on their design and study protocol, the studies were grouped into repeatability studies and reproducibility studies; repeatability studies compared measurements obtained using the same method (e.g., an ETDRS test-retest) under identical or nearly identical conditions, while reproducibility studies compared measurements obtained using two different methods (e.g., Snellen vs. Landolt C), as defined by Bartlett and Frost.^17^ In theory, the measurement error in a repeatability study should be lower compared to a reproducibility study, as it is less prone to bias. The participants were subgrouped into healthy subjects with no occular comorbidity and subjects with an ocular comorbidity; in studies that included both subgroups, each subgroup was reported separately. The results were visually reported as the mean differences and LoA in forest plots; in addition, a summarized mean of the LoA ranges is reported with 95% confidence interval (CI). To create the forest plots, the “forestplot” package (version 3.1.3) was used with R statistical software (version 4.1.2; Comprehensive R Archive Network, Vienna, Austria).

### Ethics statement

This systematic review exclusively used data obtained from previously published studies and did not include any novel research involving human participants or animals conducted by the authors. In accordance with national Dutch regulations, approval from an institutional ethics review board was not required.

## Results

### Search

Our literature search initially identified 2,427 records for VA and 2,335 records for RE. After applying the inclusion and exclusion criteria, and removing duplicate records, a total of 19 studies (13 involving VA and 6 involving RE) published in 1998 through 2022 were included in our analysis. The studies reporting on VA either compared measurements obtained using different charts (ETDRS, Snellen line and letter, Landolt C, Tumbling E, and/or Bailey-Lovie) or were test-retest assessments using the same chart.

### Quality assessment

The results of the QUADAS-2 tool used to assess RoB and applicability are reported for both VA and RE. Only 1 study scored a low RoB, while 13 studies scored a high or unclear RoB in at least three out of four QUADAS-2 domains. Most concerns were reported in the Index Test domain; for example, the test protocols were not described (or were not described sufficiently), the observers were not blinded to the outcome of the first test when performing the second test, and/or the test order was not random. For studies involving VA, RoB in the Flow and Timing domain was low in 8/13 studies (62%); In contrast, 5/6 studies (83%) involving RE refraction had high RoB in the Flow and Timing domain. RoB was high for all other QUADAS-2 domains in at least 50% of studies. All 13 VA studies scored a high rate of applicability (≥2 out of 3) and matched the research question; in contrast, only 3 of the 6 RE studies scored a high rate of applicability.

### Visual acuity measurements

A forest plot summarizing the results regarding agreement for VA is stratified for studies that reported data for subjects with no ocular comorbidity and studies that reported data for subjects with an ocular comorbidity. For the subjects with no ocular comorbidity, the structural bias (i.e., the mean difference) ranged from -0.12 to 0.07 LogMAR (summarized mean structural bias: -0.03 LogMAR; 95% CI: -0.07, 0.01), with no apparent differences between the charts used or study design (e.g., repeatability versus reproducibility). For the subjects with an ocular comorbidity, the range of structural bias was considerably wider, ranging from -0.14 to 0.32 LogMAR (summarized mean structural bias: 0.07 LogMAR; 95% CI: 0.02, 0.12). The overall summarized mean of the structural bias (i.e., for all subjects combined) was 0.03 LogMAR (95% CI: -0.01, 0.07).

Random error—expressed as the 95% LoA—ranged from ±0.10 to ±0.31 LogMAR (summarized mean random error: ±0.15 LogMAR; 95% CI: 0.12, 0.19) among the subjects with no ocular comorbidity. For subjects with an ocular comorbidity, random error ranged from ±0.10 to ±0.49 LogMAR (summarized mean random error: ±0.23 LogMAR; 95% CI: 0.18, 0.27). The overall summarized mean random error was ±0.20 LogMAR (95% CI: 0.17, 0.23). Nine of thirty subgroups among six different studies reported a random error that was actually within the clinically accepted limit of ±0.15, although three subgroups reported high structural bias of at least 0.10 LogMAR.^10,20,22,23,25,26^ Finally, the methodological quality was rates as suboptimal, as four of these six studies reported a high RoB for at least 3 out of 4 QUADAS-2 domains.

### Refractive error measurements

A forest plot summarizes the results regarding agreement for RE. The structural bias (i.e., mean difference) ranged from -0.01D to 0.60D (summarized mean structural bias: 0.14D; 95%: CI -0.02, 0.29). Random error expressed as 95% LoA ranged from ±0.49D to ±1.27D (summarized mean random error: ±0.70D (95% CI: 0.50, 0.89). Two studies reported a random error within ±0.5D; however, it should be noted that for the study by Taneri et al. the results of the first RE measurement were used as a starting point for the second RE measurement. With the exception of the study by Bullimore et al., all studies reported a high RoB in at least three QUADAS-2 domains.

### Relevant studies that did not meet the inclusion criteria

The results of any studies that did not meet the inclusion criteria for our systematic review—but were still considered relevant to the discussion—are presented in Supplementary Table S2. These six studies—two regarding VA and four regarding RE—were not included because they did not directly compare VA charts, did not report LoA or mean difference, or duplicated data from another study that was included in the analysis. Interestingly, five of these six excluded studies exceeded the clinically accepted deviation limits of random error.

Studies that compared the accuracy of measuring manifest refraction vs. an autorefractor were also excluded from our analysis, as the reported variation here is considered a proxy for the variation between repeated measurements of manifest refraction. However, for the sake of comparison we identified these papers in our literature search, and the results are summarized in Supplementary Table S3. Fifteen out of 16 such studies reported a random error that exceeded the clinically accepted deviation limits; specifically, random error ranged from 0.47D to 1.39D (summarized mean: ±0.83D; 95% CI: 0.68, 0.97), showing higher variability compared to the studies included in our analysis.

## Discussion

The goal of this systematic review was to evaluate the validity of current variability thresholds regarding acceptable variations in VA and RE based on previous studies.

This review included 19 studies, of which only 9 were found to report a level of variability within the suggested clinically accepted deviation limit of ±0.15 LogMAR for VA or ±0.5D for RE. Of these nine studies, six scored a high RoB in at least three out of four QUADAS-2 domains. Seven of these nine studies were VA studies, of which three reported a remarkably high systematic bias (from 1-3 lines of letters). Thus, the majority of studies reported either high RoB or high variability, or both.

The VA studies reported a substantial difference in terms of random error in the subgroup of subjects with an ocular comorbidity compared to subjects without an ocular comorbidity. Focusing on this considerable difference in random error—irrespective of the RoB and structural bias assessments— these results suggest that the current clinically accepted deviation limit is more feasible for use in subjects without an ocular comorbidity, but is far from ideal for use in subjects with an ocular comorbidity. This finding suggests that random error is context-dependent, and the clinically acceptable deviation limit should take this context into account. Thus, if in doubt regarding whether random error should be adjusted for a given population, we recommend using the overall summarized mean random error of ±0.20 LogMAR obtained in our review.

Overall, our results suggest that the validity of the variability cannot be judged properly due to the high RoB identified in most studies. However, even if this is ignored, evaluating the variability of summarized mean of LoA indicates that both measurements have higher variability than previously suggested. This does not align with what is currently considered a clinical accepted deviation limit for individual cases. Whether this finding should be cause for concern depends on individual clinical, regulatory, and scientific perspectives. Although the summarized means of random error may serve as a best guess for establishing a new reference point, we recommend increasing knowledge, awareness, and understanding of the variability associated with measuring VA and RE—rather than attempting to establish new deviation limits.

Three of the studies in our review had relevant limitations or exhibited irregularities that warrant discussion, and we consider that the results of these studies may not be representative. First, one study reported “95% tolerance limits” rather than 95% LoA. Although the authors suggest that this metric is interchangeable with 95% LoA, the formula they reported for their calculations raises concerns regarding whether they are indeed interchangeable. Moreover, we could not verify the exact number of comparisons across three assessed charts, as the authors reported 80 measurements in 40 subjects. The second study reported that their study included 504 subjects; however, only 28 of the 504 measurement points can be discerned in their Bland-Altman plots. While some measurement points may overlap, the exact number of measurements performed remains unclear. Moreover, the authors claimed that they “confirmed” the Snellen results using a Tumbling E chart in order to avoid false readings, before comparing the Snellen results to ETDRS results; this re-testing before comparing can introduces bias. For example, the authors did not report whether any Snellen results were omitted based on that confirmation. The authors also reported LoA values of ±0.31 LogMAR and ±0.10 LogMAR for measurements performed before and after cataract surgery, respectively. In contrast, other studies did not report a difference in variability of this magnitude due to cataract surgery. It is therefore possible that the authors compared preoperative unaided distance visual acuity (UDVA) with postoperative best corrected visual acuity (BCVA), but this is only speculation, as the authors provided only limited details regarding their postoperative study population, and we believe they did not sufficiently address the LoA difference in their discussion. Finally, the third study reported that their first RE measurement was used as a starting point for their second RE measurement, resulting in potential bias. However, the authors stated that if any discrepancies were found between the first and second measurements, a third measurement was performed, and that any measurements could be repeated “as deemed necessary, and/or surgeon’s (confirmatory) manifest refraction was performed”. This is relevant, as by definition the results obtained with repetitive measurements reduce the amount of measurement variation within a subject and therefore may not necessarily represent the real-world variation in measuring RE. Moreover, the authors failed to specify which measurements were compared, or how many third measurements were performed. Overall, the design of their study raises concerns regarding confirmation bias, and their results may potentially overestimate the agreement of RE measurements.

We also identified six studies that did not meet our inclusion criteria, but may be relevant. The study by Siderov et al.^14^ is often cited by other studies stating that ±0.15 LogMAR is reasonable. Unfortunately, we could not include this study in our analysis, as the authors did not directly compare VA charts, but interchangeably used Snellen, Bailey-Lovie, and ETDRS charts to measure VA within the same subject. Nevertheless, they reported LoA to be ±0.16 LogMAR. The study regarding RE by Rosenfield et al.^35^ is also cited often as a landmark study, yet the authors repeated their MR measurement on each subject five times and calculated compared means rather than individual measurements. Moreover, the authors did not report how their outcomes were calculated. Nevertheless, they reported an LoA of ±0.61D by comparing MR with autorefractors rather than comparing MR with MR. The study be Raasch et al.^36^ was not included in our review because their data were retrieved from another study; however, they reported an LoA of ±1.49D. Lastly, Pesudovs et al.^37^ did not report mean differences, but an examination of their data suggests these differences to be close to zero. Although the authors calculated LoA for MR across four observers (LoA for SEQ ±0.48D), they did not state how LoA was calculated (e.g., whether it was the average of four pairwise comparisons), thus making it difficult to assess the reproducibility of these results.

A frequently cited, authoritative study that warrants discussion is the 1996 review by Goss and Grosvenor^13^ in which the authors concluded that RE measurements are repeatable within a range of ±0.25D to ±0.5D. However, the studies included in their review used various methodologies, reporting standards, outcome measurements, and measurement techniques (e.g., MR, autorefractor, or retinoscopy). It should also be noted that their review was conducted before the publication of relevant guidelines for systematic reviews and RoB assessment, thus warranting a re-evaluation of the available evidence. Moreover, their methodology did not account for the differences in measurements outside of the clinical accepted deviation limit (e.g., ±0.5D), as this can be properly assessed only using 95% LoA. For example, if a study has a considerable number of measurements that fall outside of the ±0.5D limit, percentage-based metrics are minimally affected. Notably, although they used percentage-based metrics, a considerable number of studies failed to report that 95% of measurements fell within the ±0.5D limit, calling into question the authors’ conclusion that measuring RE is consistently repeatable within a range of ±0.25D to ±0.5D; indeed, we believe that this is not the case.

Another noteworthy study was published by Shah et al. 2009, in which they examined RE measurements.^38^ Although having a similar design as the studies by two of our included studies, this study by Shah et al. did not meet our inclusion criteria because they did not report relevant outcomes. In their study, 102 randomly selected optometrists examined three standardized patients, and the authors reported reproducibility ranging from 0.5D to 0.75D, demonstrating that even with trained patients, refraction measurements can vary by up to 0.75D between observers. This finding underscores the inherent variability when measuring RE and is consistent with the results reported in our review. One possible explanation for this inherent variability is that MR is guided by the VA result achieved with the prescription, thus likely reflecting both the variance in VA testing and the RE measurement itself.

### Strengths and limitations

This is the first systematic review regarding the clinically accepted deviation limits of both VA and RE. This review broadens the scope of the field by including the most commonly used charts for measuring VA (namely, the ETDRS, Snellen, Bailey-Love, Landolt C, and Tumbling E charts). These charts are highly relevant to RE, as VA is often used to guide and refine RE measurements. On the other hand, this review has some limitations that warrant discussion. First, with respect to RE we included only studies that compared MR measurements, even though autorefractors are also frequently compared and are potentially relevant in the field of RE. Nevertheless, MR is considered the gold standard for measuring RE, and by using only one outcome measure we attempted to ensure that our analysis is both clear and cohesive. However, it is worth mentioning that much can be learned from objectively measuring RE, and results obtained with an autorefractor are often used as a starting point for measuring MR. For this reason, we included the studies identified in our search that compared autorefractors with MR as a frame of reference. Second, assessing RoB is a subjective process, as researchers may prioritize specific aspects as being important and/or relevant, and answers can vary, as they are based on the reader’s interpretation. However, we believe that these assessed risk elements play an essential role in appropriately interpreting the results of our review. Third, we believe that blinding of the results—in which the test-taker does not know the outcome of the first measurement before the second measurement is taken—to be relevant for making appropriate comparisons. Although we understand this is a challenge in a repeatability design, with only one test-taker allowed, we still encourage researchers to use a blinded approach, a concept that is fundamental to solid, reliable research in general.

### Methodological recommendations

Our systematic review demonstrates that measuring both VA and RE has higher inherent variability than previously realized, while also highlighting both limitations and inconsistencies in the literature. To provide more accurate estimates of the repeatability and reproducibility of these measurements, we believe that more methodologically rigorous and comparable studies are needed. Based on our findings, we offer several recommendations for future research, particularly regarding the methodological practices used and the reporting of results.

For repeatability studies, both measurements should be performed under identical—or as near to identical as possible—conditions, ideally on the same day, while still accounting for the potential effects of fatigue due to multiple measurements. This is not necessarily a strict requirement for reproducibility, as variables will vary naturally by study design and may better reflect real-world variation. However, researchers should also consider—and, if necessary, examine—the effects of confounding variables such as environmental factors that may affect agreement between subsequent measurements.

A common issue in many studies involving RE is the recruitment of participants from optometry schools and/or frequent testing of the same subjects (for example, as many as 40 times), which can limit the generalizability of the findings, particularly given that these subjects are not representative of the general population. Therefore, we recommend including naïve patients in these studies wherever possible.

Blinding the observers to the results of previous measurement is recommended as a best practice. If blinding is not feasible or practical, randomizing the observers and/or the test sequence is the next best option. Another best practice is to estimate agreement using the Bland-Altman method, reporting both structural bias and random error, ideally combined with the 95% confidence interval for both. In addition, re-sampling methods such as bootstrapping should be used to estimate population means, particularly when both of the subjects’ eyes are included in the analysis.

### Conclusions

Our results underscore the notion that both VA and RE measurements are inherently variable. Moreover, the current clinically accepted limits of this variation are either not supported by the majority of studies in our review or are accompanied by a relatively high risk of bias, particularly among subjects with an ocular comorbidity. Thus, future studies using high methodological rigor are needed in order to better estimate more accurate limits of variability. Until then, we propose that the overall summarized mean of random error identified by our review—namely, ±0.20 LogMAR and ±0.70D for VA and RE, respectively—should be used as a provisional frame of reference. Moreover, our review provides several teaching opportunities with respect to understanding variability among measurements and the methodological challenges associated with designing these studies.

## Data Availability

All data produced in the present study are available upon reasonable request to the authors

## Acknowledgements

None.

## Conflict of Interest

RW is a shareholder and advisor at Easee BV; MM is an employee at Easee BV. The authors have no additional financial or proprietary interest in the materials presented herein.

## Financial Support

None

## Ethics Approval

Analyses were performed in accordance with Dutch privacy laws and the Declaration of Helsinki in the context of quality control and health care evaluation. In accordance with the Dutch Central Committee on Research Involving Human Subjects (CCMO) guidelines, ethics approval and informed consent were not required.

## Abbreviations

D: Diopters
LoA: Limits of Agreement
MR: Manifest Refraction
VA: Visual Acuity
CDVA: Corrected Distance Visual Acuity
UDVA: Unaided Distance Visual Acuity
BCVA: Best Corrected Visual Acuity
LogMAR: Logarithm of the Minimum Angle of Resolution
ETDRS: Early Treatment Diabetic Retinopathy Study
ODS: Oculus Dexter Sinister (right and left eye)
PRISMA: Preferred Reporting Items for Systematic Reviews and Meta-Analyses
PROSPERO: International Prospective Register of Systematic Reviews
SD: Standard Deviation
RoB: Risk of Bias

## Figure legends

**Figure 1.**
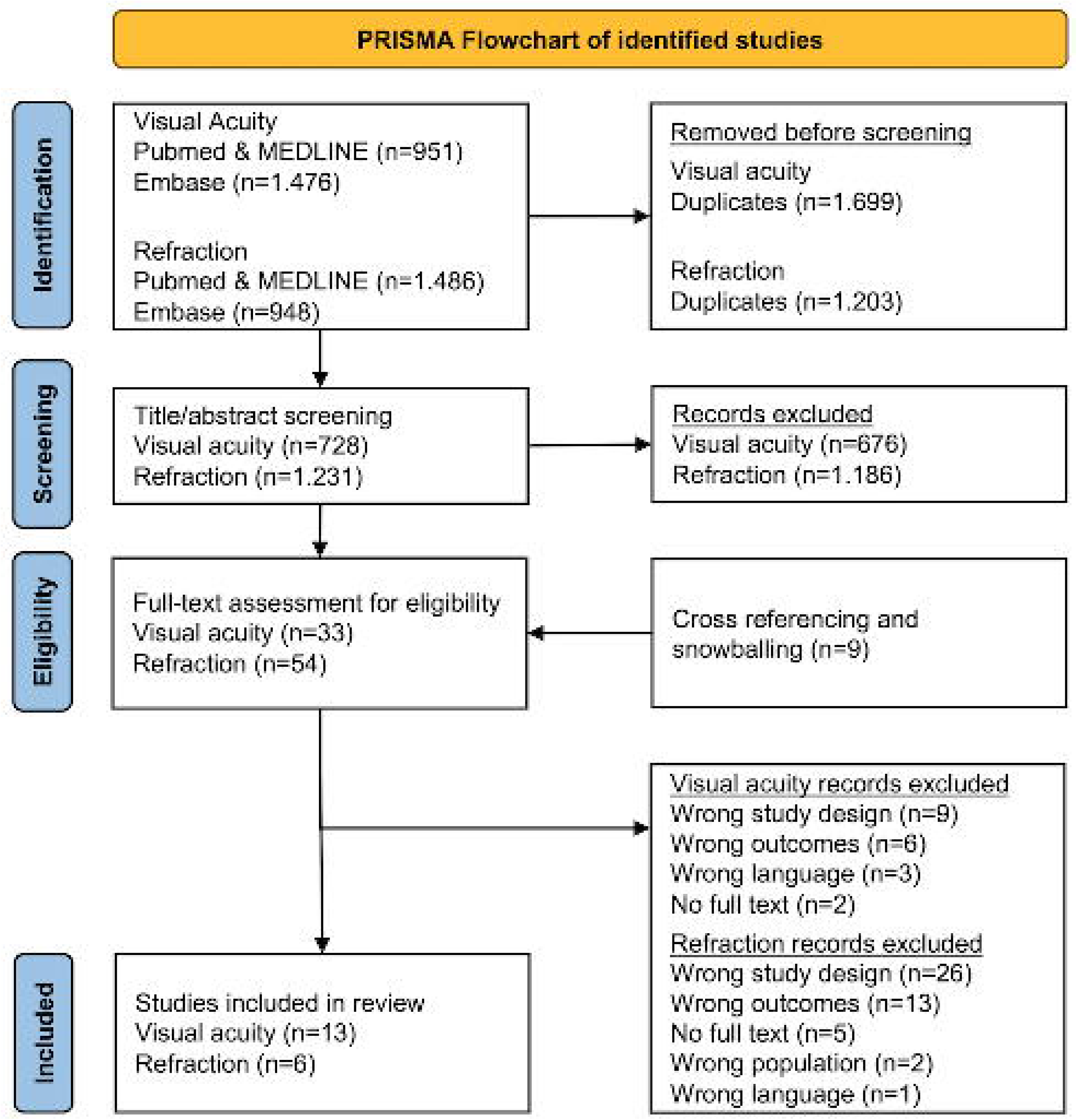
Flow chart detailing the identification, screening, eligibility, exclusion, and final studies included in our analysis. The studies were identified in accordance with the Preferred Reporting Items for a Systematic Reviews and Meta-Analyses (PRISMA) guidelines.

**Figure 2.**
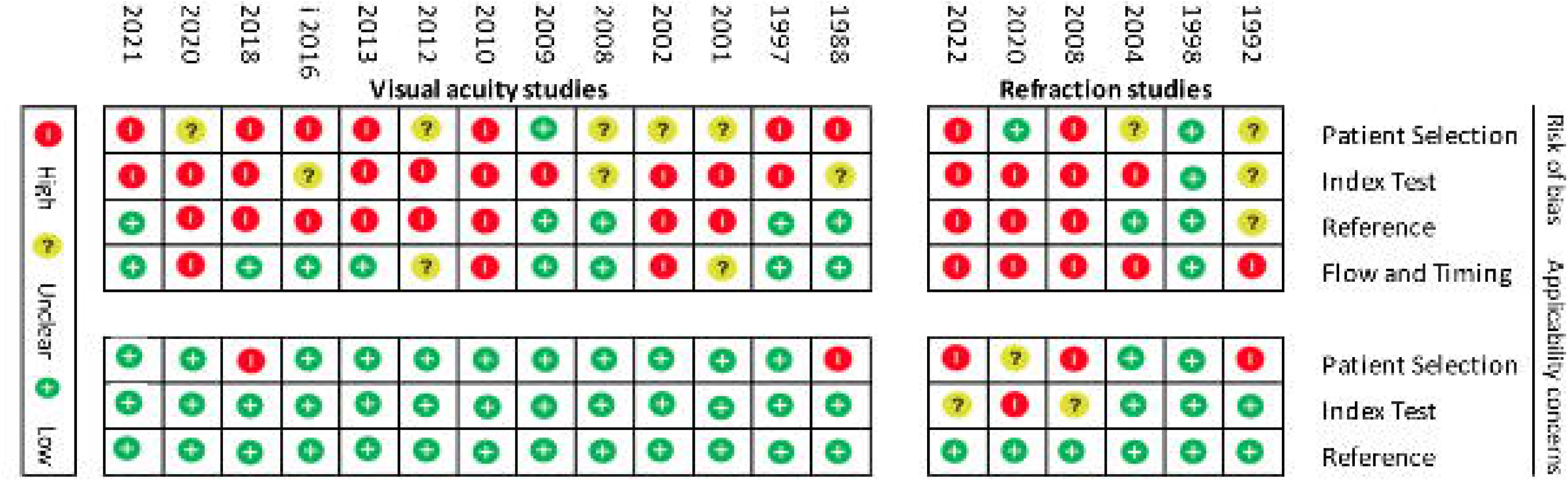
Risk of bias assessment based on the QUADAS-2 tool for studies involving visual acuity (n=13) and studies involving refractive error (n=6).^16^ Also shown are the applicability concerns for all 19 studies.

**Figure 3.**
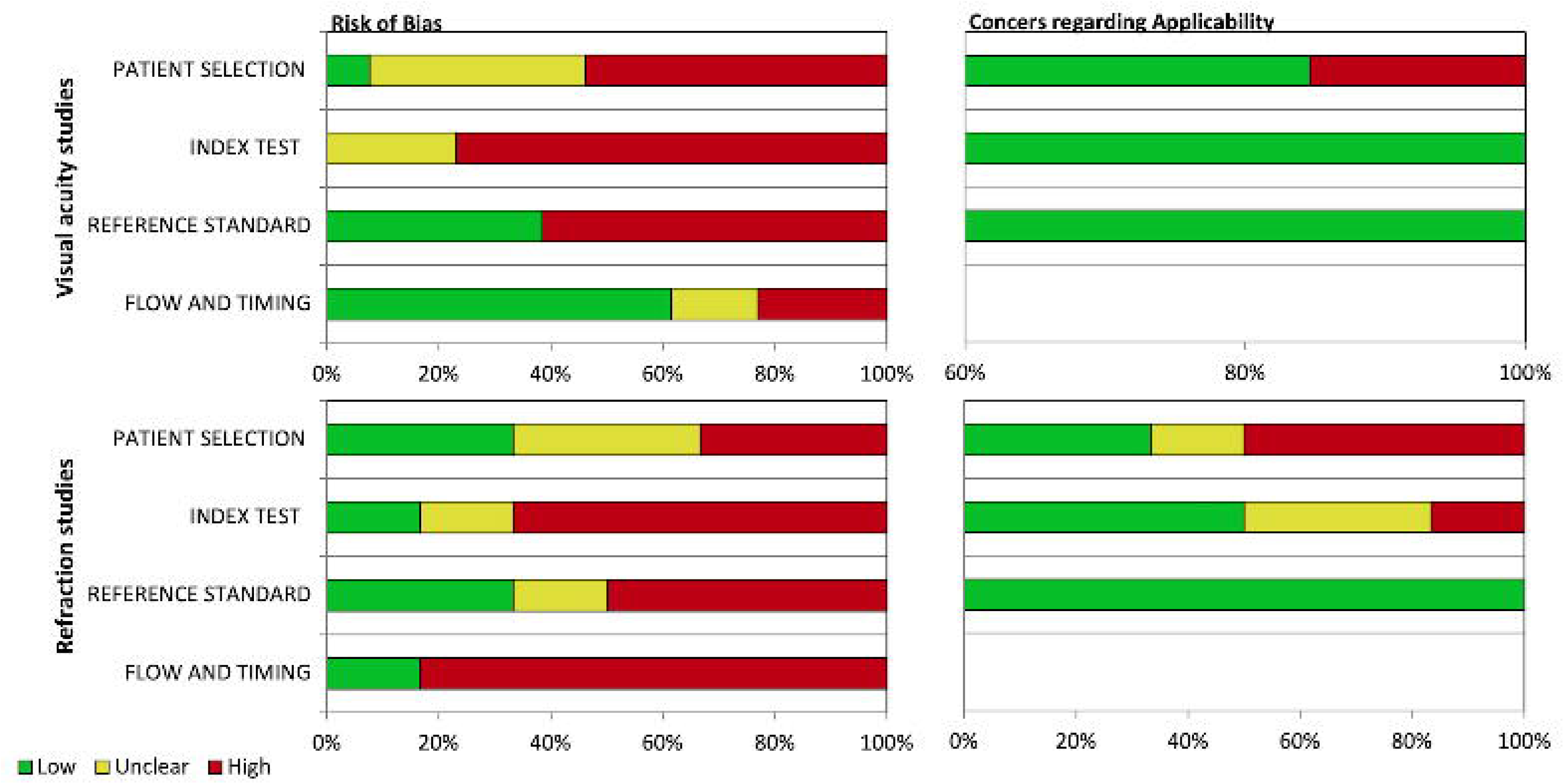
Graphical representation of the risk of bias assessment and applicability concerns for the VA and RE studies, based on the QUADAS-2 tool.^16^

**Figure 4.**
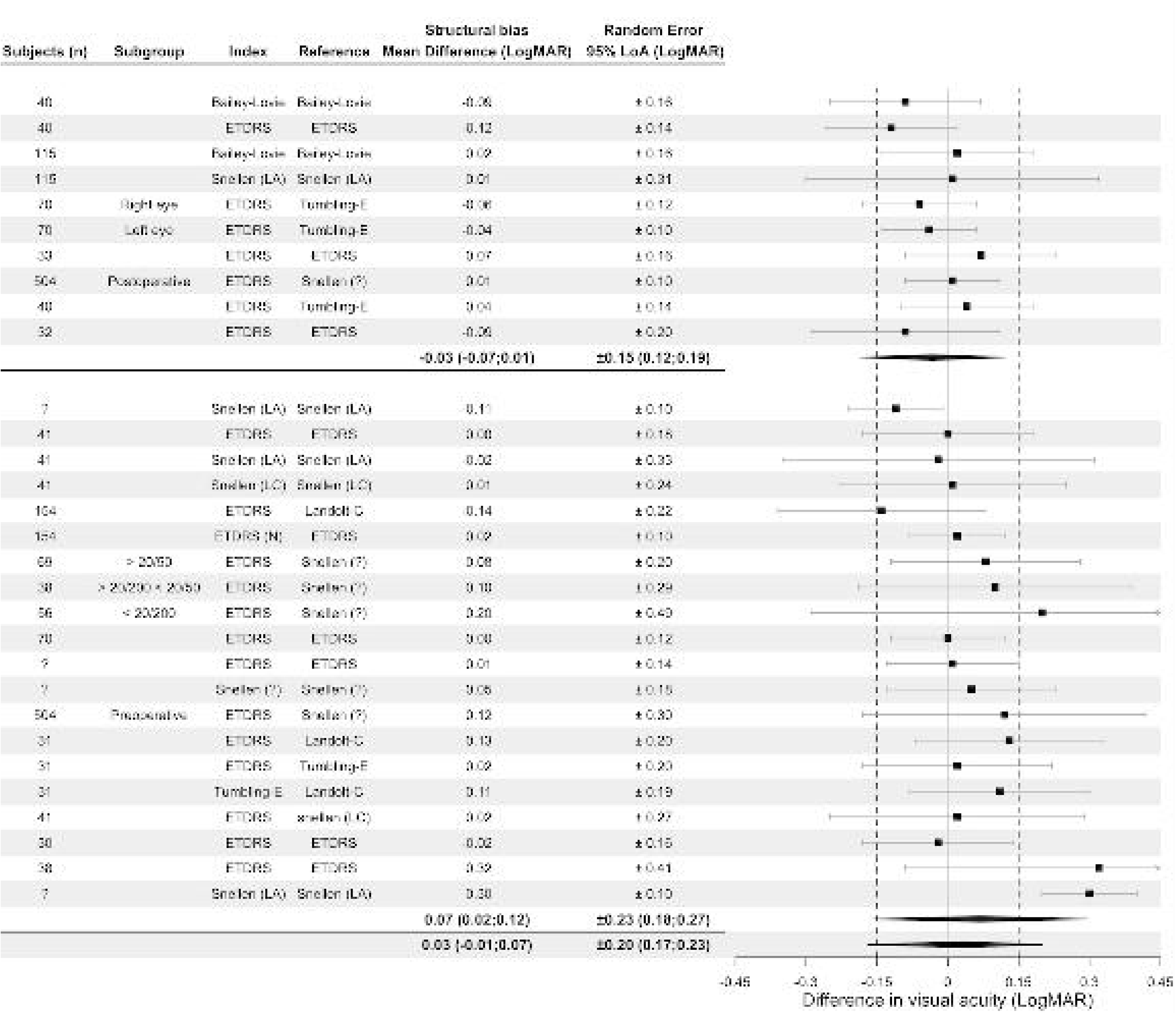
Forest plot summarizing the studies that assessed VA, stratified by patients with on ocular comorbidity (top) and patients with an ocular comorbidity (bottom). The vertical dashed lines indicate ±0.15 LogMAR. Studies marked with the † symbol are repeatability studies; all other studies are reproducibility studies. Studies marked with an asterisk (*) have a high risk of bias. LA, line assignment; LC, letter count; N, numerical optotypes; ETDRS, Early Treatment Diabetic Retinopathy Study; LoA, 95% limits of agreement; ?, not reported; 95% CI, 95% confidence interval; LogMAR, logarithm of the minimum angle of resolution; VA, visual acuity.

**Figure 5.**
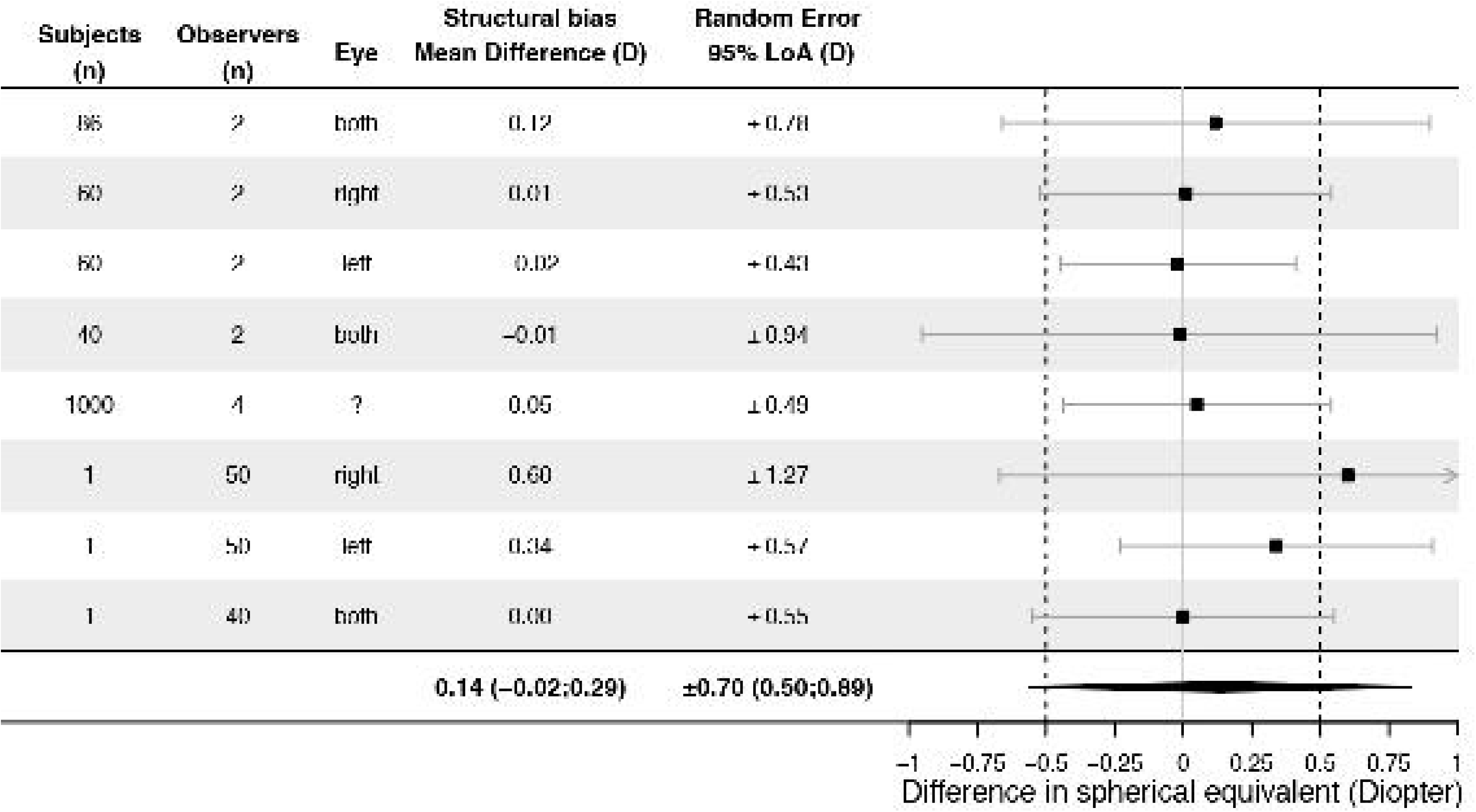
Forest plot summarizing the studies that assessed RE using manifest refraction. The vertical dashed lines represent ±0.5 D. Studies marked with an asterisk (*) have a high risk of bias. LoA, 95% limits of agreement; ?, not reported; D, diopters; RE, refraction error; 95% CI, 95% confidence interval.

## Figures

### Clarifying Methodological Concepts

#### Confidence interval vs. limits of agreement

Both the 95% confidence interval (CI) and the 95% limits of agreement (LoA) are expressed as a range, but they serve different purposes. Unfortunately, 95% CI and 95% LoA are often used interchangeably. CI indicates the precision of estimated bias between two methods, in our case regarding the mean difference or LoA; specifically, it represents the range within which the true value of the mean difference of LoA is likely to fall, thereby estimating the precision of a given value. In contrast, LoA reflects the range within which 95% of individual differences between measurements, thereby indicating the degree of agreement between two methods. In summary, while CI reflects the precision of a value, LoA reflects the range of all measurements.

#### Structural bias vs. random error

The mean difference and LoA express the concepts of structural bias and random error, both of which are essential for interpreting the degree of agreement between two methods. Structural bias refers to the systematic difference between two methods (i.e., the mean difference) and indicates whether one method systematically overestimates or underestimates the other method. Conversely, random error refers to the variability in individual measurements, expressed here as the 95% LoA. Unlike structural bias, random error is affected by unpredictable factors. Because LoA reflects the variability of individual measurements, it reflects the range within which most (i.e., 95%) individual differences are expected to fall. In summary, structural bias assesses systematic differences, whereas random error assesses unpredictable factors.

## Tables

**Table 1.**
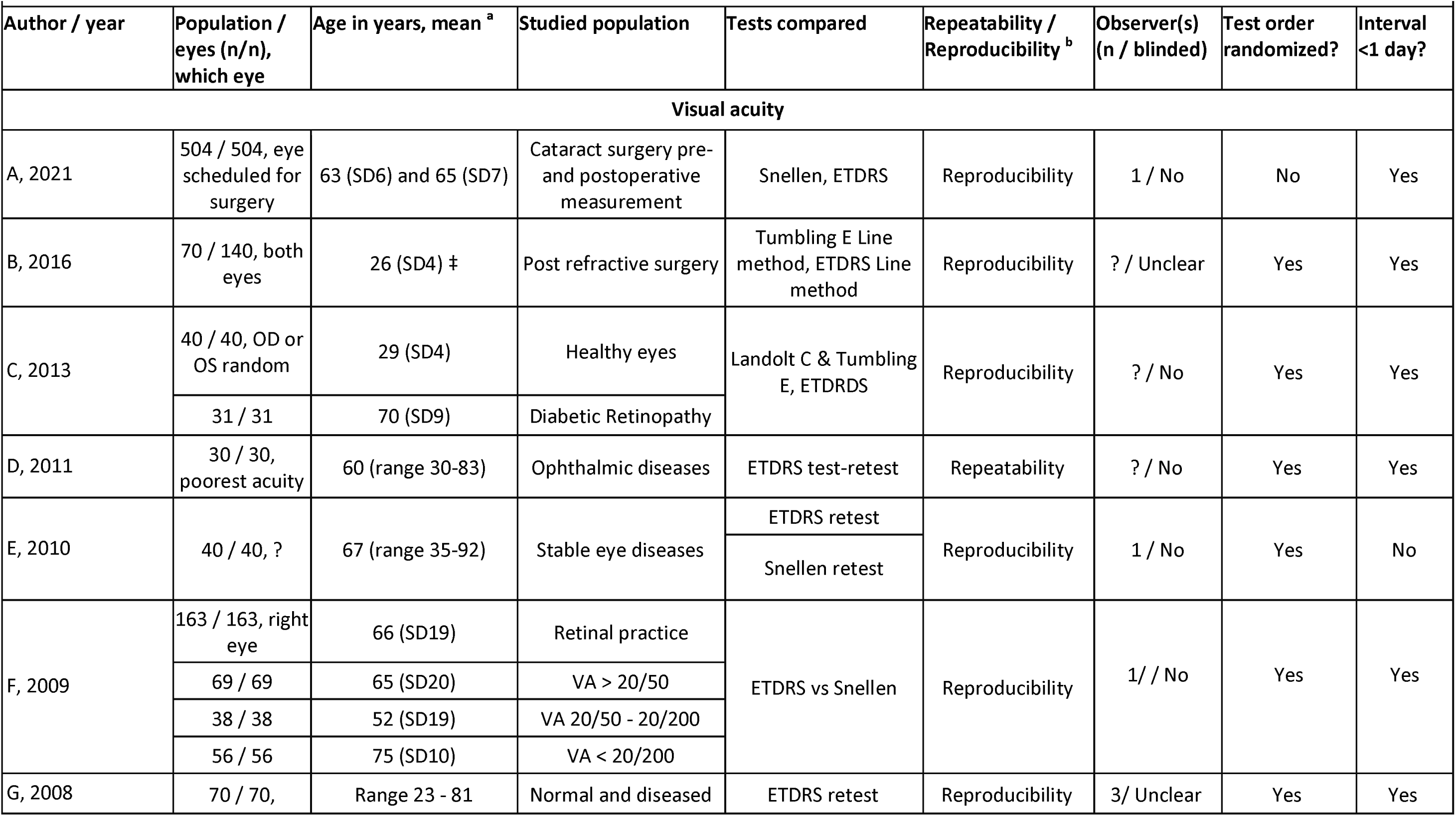
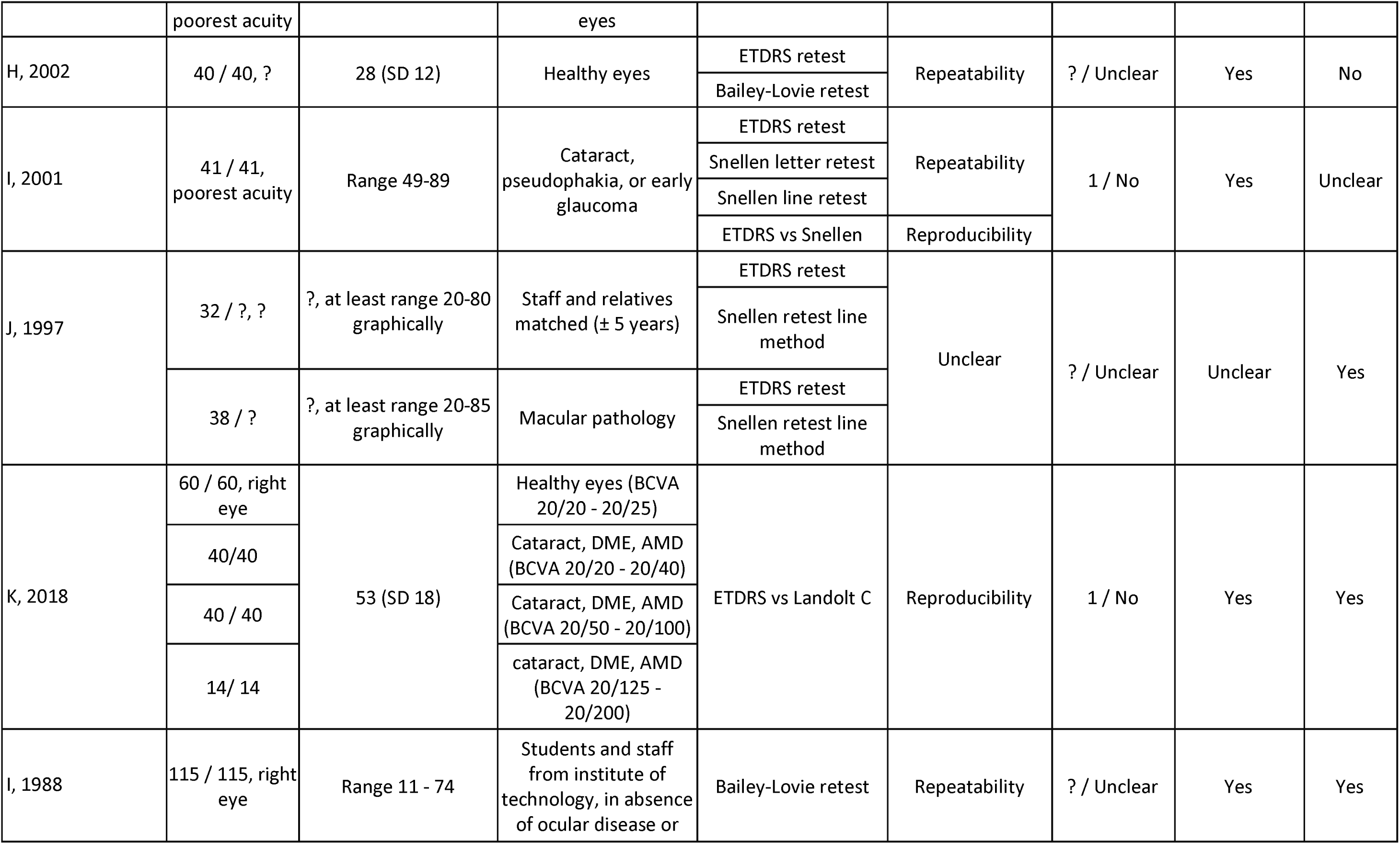
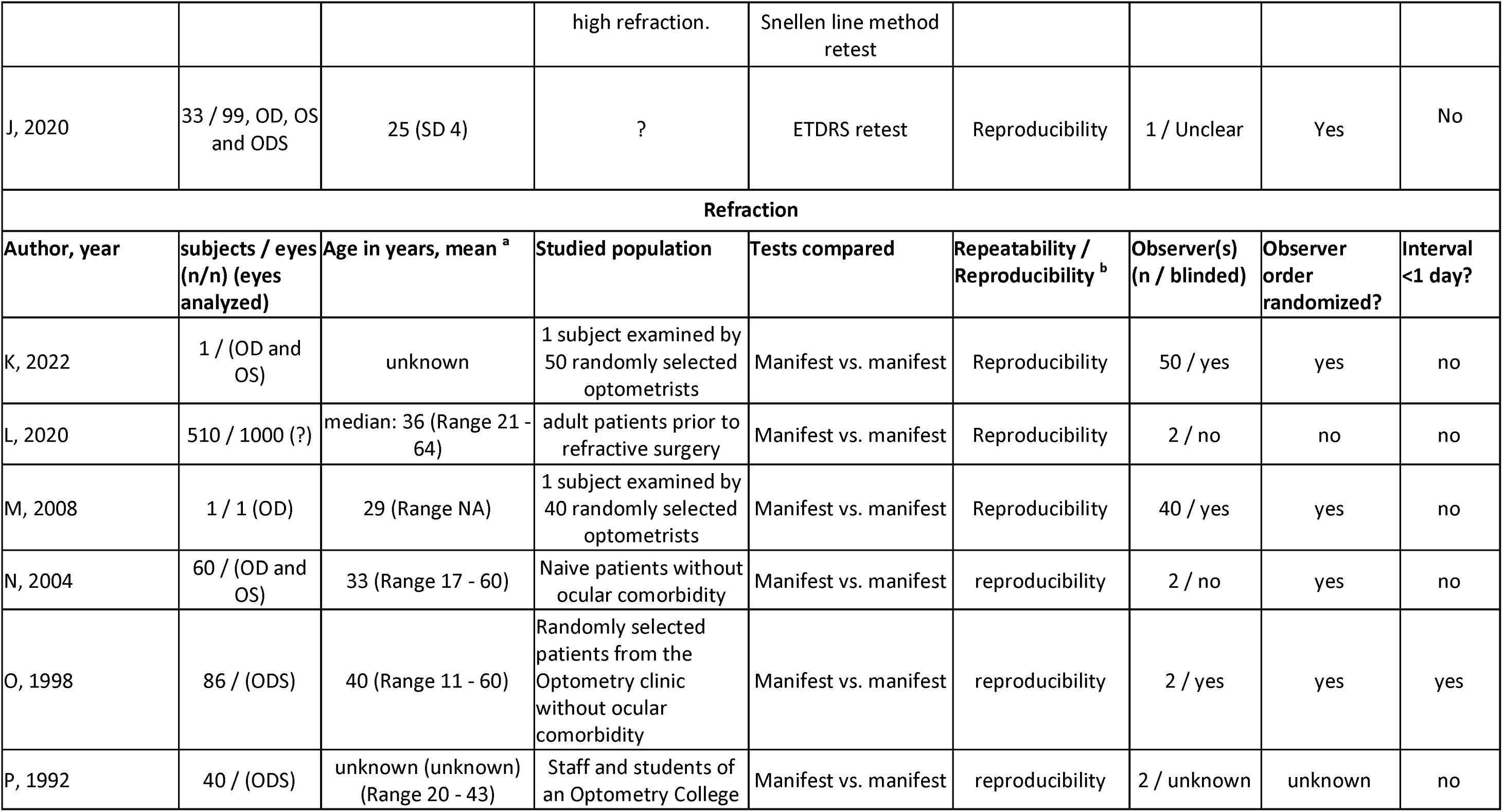
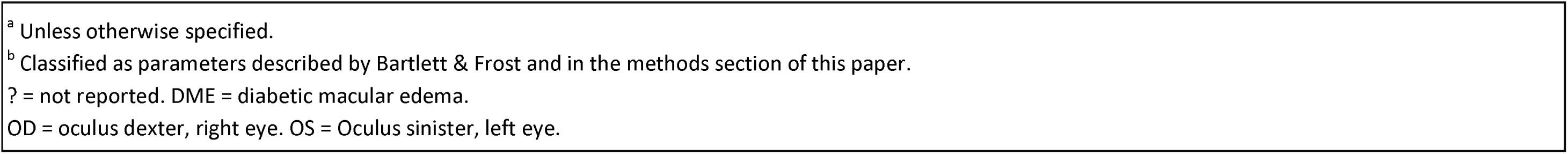
Characteristics of included studies for visual acuity and refraction

|  |  |  |  |  |  |  |  |  |
| --- | --- | --- | --- | --- | --- | --- | --- | --- |
| Author / year | Population / eyes (n/n), which eye | Age in years, mean <sup>a</sup> | Studied population | Tests compared | Repeatability / Reproducibility <sup>b</sup> | Observer(s) (n / blinded) | Test order randomized? | Interval <1 day? |
| Visual acuity |  |  |  |  |  |  |  |  |
| A, 2021 | 504 / 504, eye scheduled for surgery | 63 (SD6) and 65 (SD7) | Cataract surgery pre- and postoperative measurement | Snellen, ETDRS | Reproducibility | 1 / No | No | Yes |
| B, 2016 | 70 / 140, both eyes | 26 (SD4) ‡ | Post refractive surgery | Tumbling E Line method, ETDRS Line method | Reproducibility | ? / Unclear | Yes | Yes |
| C, 2013 | 40 / 40, OD or OS random | 29 (SD4) | Healthy eyes | Landolt C & Tumbling E, ETDRS | Reproducibility | ? / No | Yes | Yes |
|  | 31 / 31 | 70 (SD9) | Diabetic Retinopathy |  |  |  |  |  |
| D, 2011 | 30 / 30, poorest acuity | 60 (range 30-83) | Ophthalmic diseases | ETDRS test-retest | Repeatability | ? / No | Yes | Yes |
| E, 2010 | 40 / 40, ? | 67 (range 35-92) | Stable eye diseases | ETDRS retest | Reproducibility | 1 / No | Yes | No |
|  |  |  |  | Snellen retest |  |  |  |  |
| F, 2009 | 163 / 163, right eye | 66 (SD19) | Retinal practice | ETDRS vs Snellen | Reproducibility | 1/ / No | Yes | Yes |
|  | 69 / 69 | 65 (SD20) | VA > 20/50 |  |  |  |  |  |
|  | 38 / 38 | 52 (SD19) | VA 20/50 - 20/200 |  |  |  |  |  |
|  | 56 / 56 | 75 (SD10) | VA < 20/200 |  |  |  |  |  |
| G, 2008 | 70 / 70, | Range 23 - 81 | Normal and diseased | ETDRS retest | Reproducibility | 3/ Unclear | Yes | Yes |

|  | poorest acuity |  | eyes |  |  |  |  |  |
| --- | --- | --- | --- | --- | --- | --- | --- | --- |
| H, 2002 | 40 / 40, ? | 28 (SD 12) | Healthy eyes | ETDRS retest | Repeatability | ? / Unclear | Yes | No |
|  |  |  |  | Bailey-Lovie retest |  |  |  |  |
| I, 2001 | 41 / 41, poorest acuity | Range 49-89 | Cataract, pseudophakia, or early glaucoma | ETDRS retest | Repeatability | 1 / No | Yes | Unclear |
|  |  |  |  | Snellen letter retest |  |  |  |  |
|  |  |  |  | Snellen line retest |  |  |  |  |
|  |  |  |  | ETDRS vs Snellen | Reproducibility |  |  |  |
| J, 1997 | 32 / ?, ? | ?, at least range 20-80 graphically | Staff and relatives matched (± 5 years) | ETDRS retest | Unclear | ? / Unclear | Unclear | Yes |
|  |  |  |  | Snellen retest line method |  |  |  |  |
|  | 38 / ? | ?, at least range 20-85 graphically | Macular pathology | ETDRS retest |  |  |  |  |
|  |  |  |  | Snellen retest line method |  |  |  |  |
| K, 2018 | 60 / 60, right eye | 53 (SD 18) | Healthy eyes (BCVA 20/20 - 20/25) | ETDRS vs Landolt C | Reproducibility | 1 / No | Yes | Yes |
|  | 40/40 |  | Cataract, DME, AMD (BCVA 20/20 - 20/40) |  |  |  |  |  |
|  | 40 / 40 |  | Cataract, DME, AMD (BCVA 20/50 - 20/100) |  |  |  |  |  |
|  | 14/ 14 |  | cataract, DME, AMD (BCVA 20/125 - 20/200) |  |  |  |  |  |
| L, 1988 | 115 / 115, right eye | Range 11 - 74 | Students and staff from institute of technology, in absence of ocular disease or | Bailey-Lovie retest | Repeatability | ? / Unclear | Yes | Yes |
|  |  |  | high refraction. | Snellen line method retest |  |  |  |  |
| J, 2020 | 33 / 99, OD, OS and ODS | 25 (SD 4) | ? | ETDRS retest | Reproducibility | 1 / Unclear | Yes | No |
| **Refraction** |  |  |  |  |  |  |  |  |
| **Author, year** | **subjects / eyes (n/n) (eyes analyzed)** | **Age in years, mean <sup>a</sup>** | **Studied population** | **Tests compared** | **Repeatability / Reproducibility <sup>b</sup>** | **Observer(s) (n / blinded)** | **Observer order randomized?** | **Interval <1 day?** |
| K, 2022 | 1 / (OD and OS) | unknown | 1 subject examined by 50 randomly selected optometrists | Manifest vs. manifest | Reproducibility | 50 / yes | yes | no |
| L, 2020 | 510 / 1000 (?) | median: 36 (Range 21 - 64) | adult patients prior to refractive surgery | Manifest vs. manifest | Reproducibility | 2 / no | no | no |
| M, 2008 | 1 / 1 (OD) | 29 (Range NA) | 1 subject examined by 40 randomly selected optometrists | Manifest vs. manifest | Reproducibility | 40 / yes | yes | no |
| N, 2004 | 60 / (OD and OS) | 33 (Range 17 - 60) | Naive patients without ocular comorbidity | Manifest vs. manifest | reproducibility | 2 / no | yes | no |
| O, 1998 | 86 / (ODS) | 40 (Range 11 - 60) | Randomly selected patients from the Optometry clinic without ocular comorbidity | Manifest vs. manifest | reproducibility | 2 / yes | yes | yes |
| P, 1992 | 40 / (ODS) | unknown (unknown) (Range 20 - 43) | Staff and students of an Optometry College | Manifest vs. manifest | reproducibility | 2 / unknown | unknown | no |

<sup>a</sup> Unless otherwise specified.
<sup>b</sup> Classified as parameters described by Bartlett & Frost and in the methods section of this paper.
? = not reported. DME = diabetic macular edema.
OD = oculus dexter, right eye. OS = Oculus sinister, left eye.

**Supplementary table 1.** Search Strategy.

| Supplementary table 1. Search Strategy. |
| --- |
| Search visual acuity |
| <u>Patient terms</u><br>Adult*<br>Aged<br>Middle-aged<br>Aged<br>Elderly<br>Adulthood |
| <u>Intervention terms</u><br>Visual acuity chart<br>Visual acuity test<br>ETDRS chart<br>Snellen chart<br>Landolt-C chart<br>Tumbling-E chart |
| Search refraction |
| <u>Intervention terms</u><br>Refraction AND measurement*<br>Manifest refraction AND measurement*<br>Spherical equivalent refraction AND measurement* |

**Supplementary table 2.** Visual acuity of manifest refraction studies that did not meet inclusion criteria, yet were considered relevant by other studies.

| Author | year | Subjects (n) | Mean bias (SD) | 95% LoA |
| --- | --- | --- | --- | --- |
| Q | 1999 | 50 | -0.05 (?) LogMAR | ±0.16 LogMAR |
|  |  | 25 | -0.04 (?) LogMAR | ±0.16 LogMAR |
| R † | 1993 | 5 (highly practiced healthy subjects, performing 156 retests) | ? (?) | ? |
| S † | 1996 | 17 | ? (?) | ? |
| T | 2009 | 100 | ? (?) | ±0.55 D |
| U | 2001 | 40 | 0.03 (0.51) D | ±1.49 D |
| V | 2007 | 60 | ? (?) | ±0.48 D |

D = Diopters. ? = Not Reported. LogMAR = Logarithm of the Minimum Angle of Resolution. Studies marked with † did not use Bland Altman analysis, which is considered the gold-standard for assessing agreement between two measurements.

**Supplementary table 3.** Autorefractor reproducibility (autorefractor versus manifest refraction)

|  |  |  |  |  |
| --- | --- | --- | --- | --- |
| Author | year | Subjects (n) | Structural bias<br>Mean Difference<br>(SD) | Random Error<br>95% LoA (D) |
| W,* | 1992 | 40 | -0.16 (0.56) | ±1.10 |
| X | 1995 | 12 | ? (?) | ±0.61 |
| Y | 1996 | 224 | -0.23 (0.47) | ±0.92 |
|  |  | 224 | -0.2 (0.46) | ±0.90 |
|  |  | 80 | -0.13 (0.27) | ±0.53 |
|  |  | 80 | -0.11 (0.26) | ±0.51 |
| Z* | 1998 | 86 | -0.07 (0.42) | ±0.82 |
| AA | 2002 | 20 | 0.18 (0.71) | ±1.39 |
| BB | 2005 | 192 | -0.44 (0.54) | ±1.06 |
| CC | 2009 | 50 | 0.33 (0.55) | ±1.08 |
| DD | 2015 | 243 | 0.54 (0.57) | ±1.12 |
| EE | 2015 | 60 | 0.00 (0.26) | ±0.51 |
| FF | 2018 | 1238 | -0.07 (0.24) | ±0.47 |
| GG | 2019 | 67 | -0.05 (0.38) | ±0.75 |
| HH | 2021 | 253 eyes | 0.18 (0.34) | ±0.67 |
| II | 2021 | 400 | 0.07 (0.39) | ±0.76 |
| Summarized Mean |  |  | -0.01<br>(95%CI -0.14;0.12) | ±0.83<br>(95%CI 0.68;0.97) |
| ? = Not Reported. LoA = Limits of Agreement. D = Diopters.<br>* This study reported both manifest and autorefractor data. |  |  |  |  |

## Notes

### Clinical Protocols

https://www.crd.york.ac.uk/prospero/display_record.php?RecordID=554663

### Funding Statement

This study did not receive any funding

